# Rising Tuberculosis Presentations in Ontario, Canada Are Not Explained by Community SARS-CoV-2 Activity: A Population-Based Time-Series Study

**DOI:** 10.64898/2026.11.14.26367737

**Authors:** Natalie J. Wilson, Ashleigh R. Tuite, Clara Eunyoung Lee, Rachel J. Berkowitz, Benjamin Wiebe, Jade Hall, Dagmara Boushy, David N. Fisman

## Abstract

**Background:** Tuberculosis (TB) notifications have increased in several high-income countries since 2020. Proposed explanations include pandemic-related disruption of treatment for latent tuberculosis, changes in population composition, and immune dysfunction following SARS-CoV-2 infection. We assessed whether TB presentations increased during the pandemic era and then tested whether community SARS-CoV-2 activity was followed by increased mycobacterial disease.

**Methods:** We conducted a population-based ecological time-series study of first TB- and nontuberculous mycobacterial (NTM)-coded hospital, emergency-department and ambulatory-clinic presentations in the Greater Toronto–Hamilton Area Plus (GTHA+), Ontario (2024 population approximately 11.2 million), from April 2011 through March 2024. NTM served as a comparator because it shares diagnostic pathways with TB but differs epidemiologically. Negative-binomial projection and full-series models estimated pandemic-era changes under three alternative trend assumptions. Distributed lag non-linear models estimated cumulative rate ratios for standardized community viral activity over 0–12 weeks; sensitivity analyses extended the maximum lag to 26 and 52 weeks. Influenza and respiratory syncytial virus (RSV) were prespecified comparators; data for five additional viral exposures were obtained after the primary analysis was completed and analysed as exploratory comparisons.

**Results:** We identified 3,985 first TB-coded and 5,214 first NTM-coded acute-care presentations. Across six models, estimated pandemic-era TB increases ranged from 3% to 22%. No model suggested an increased in NTM. SARS-CoV-2 activity was not positively associated with TB (cumulative rate ratio per 1-SD increase over 0–12 weeks 0.909, 95% CI 0.822–1.006) or NTM (0.953, 0.868–1.047). No positive associations were identified in subgroup analyses or with maximum lags of 26 or 52 weeks. Influenza was not associated with either outcome. RSV was associated with TB and NTM; only the RSV–NTM association met the Bonferroni threshold among the prespecified comparisons, and no exploratory associations met the experiment-wide threshold.

**Conclusions:** We identified a modest pandemic-era increase in TB-coded acute-care presentations in Ontario, Canada, although its estimated size depended on assumptions about underlying trends. NTM-coded presentations did not increase. Community SARS-CoV-2 activity was not positively associated with subsequent TB or NTM presentations at lags of up to one year and did not provide a temporal explanation for the population-level TB increase. Individual-level effects cannot be excluded.

## INTRODUCTION

Tuberculosis (TB) notifications have increased since 2020 in several high-income countries, including the United States, United Kingdom, and Canada (1–6); the Canadian incidence rate rose from 5.1 per 100,000 population in 2019 to 6.1 per 100,000 in 2024, corresponding to an increase from 1,921 to 2,508 reported cases (3–5). Several mechanisms could produce a SARS-CoV-2 pandemic-era increase in TB, including disruption of TB prevention and diagnostic services (7, 8), delayed diagnosis, changes in population composition and healthcare use, and immune effects following SARS-CoV-2 infection. Acquired immunological abnormalities have been reported following SARS-CoV-2 infection (9–14). As control of latent *Mycobacterium tuberculosis* infection depends heavily on cell-mediated immunity, post-infectious immune dysfunction could plausibly have contributed to observed pandemic-era increases in TB.

Determining whether TB presentations exceeded expectations during the pandemic requires careful evaluation of pre-pandemic trends and seasonality. However, identifying an increase beyond expectations does not, in itself, establish its cause. If such an increase resulted from immune effects of SARS-CoV-2 infection, surges in SARS-CoV-2 activity would be expected to be followed by increased TB presentations. By contrast, service disruption, migration, changes in healthcare use, or ascertainment could alter the pandemic-era level without tracking short-term viral activity. Nontuberculous mycobacterial (NTM) disease shares laboratory diagnostic pathways with TB but has different epidemiological and immunological determinants (15–18). NTM are environmental organisms acquired from water and soil rather than from other people, so that NTM disease does not propagate through transmission chains and is not subject to the contact tracing and preventive therapy that shape TB epidemiology. Pulmonary disease predominates and presents indolently, with chronic cough, sputum production and weight loss developing over months. Risk is governed largely by airway architecture, with bronchiectasis and chronic obstructive pulmonary disease the dominant predisposing conditions and a substantial burden among older women with structural lung disease (15–17); cell-mediated immune competence, which is central to control of latent M. tuberculosis infection, is a far less important determinant outside rare syndromes of Mendelian susceptibility to mycobacterial disease (15–18). NTM presentations therefore provide a useful comparator, and convergence or divergence in secular trends and associations with viral activity may be informative.

Using population-based administrative data for approximately 11 million Ontario residents over 13 years, we evaluated whether first TB- and NTM-coded acute-care presentations departed from pre-pandemic trajectories. We then tested whether weekly SARS-CoV-2 activity predicted subsequent presentations over maximum lag windows of 12, 26, and 52 weeks; whether associations differed by age or recorded immunocompromising condition; and how the results compared with those for other respiratory viruses.

## METHODS

### Study design and setting

We conducted a population-based ecological time-series study of first tuberculosis (TB)- and nontuberculous mycobacterial (NTM)-coded health care presentations (hospital admissions, emergency-department [ED] visits and visits to selected ambulatory clinics) in the Greater Toronto–Hamilton Area Plus (GTHA+), Ontario. The catchment comprised 14 public health units covering approximately 5% of Ontario’s land area and had a combined 2024 population of approximately 11.2 million, representing about 70% of Ontario’s and 27% of Canada’s populations (**Supplementary Appendix**). The outcome series extended from April 1, 2011, through March 31, 2024 (676 analyzed weeks). The 468 weeks through March 14, 2020, constituted the pre-pandemic period; the pandemic era began March 15, 2020, with the onset of community SARS-CoV-2 transmission in Ontario. For lagged analyses, observations preceding the analytic period were used only to construct exposure histories and did not contribute outcome observations.

### Case ascertainment

We ascertained weekly counts of first TB- and NTM-coded acute-care presentations from two linked Canadian Institute for Health Information (CIHI) administrative databases: the Discharge Abstract Database for acute inpatient admissions and the National Ambulatory Care Reporting System for emergency-department visits and for visits to selected ambulatory clinics, including family medicine clinics. TB was defined by ICD-10-CA codes A15, A17, A18, or A19 in any diagnosis field; A16, which denotes clinically suspected but unconfirmed pulmonary TB, was excluded as nonspecific. NTM disease was defined by code A31. We linked records using encrypted health-card numbers and retained the earliest observed qualifying record for each person separately for TB and NTM; a person could therefore contribute once to each outcome. This yielded 3,985 first TB-coded and 5,214 first NTM-coded presentations. Because these outcomes were administrative health care records rather than validated public-health notifications, and because the ambulatory component is not restricted to unscheduled care, we describe them as health care presentations rather than incident cases. Recorded immunocompromising host conditions (ICH) comprised HIV/AIDS (B20–B24, Z21), solid-organ transplantation (Z940–Z944), stem-cell or marrow transplantation (Z941, Z948), haematological malignancy (C81–C85, C88, C90–C96), or primary immunodeficiency (D80–D84). Medication-induced immunosuppression was not reliably captured; some affected people could therefore have been classified as not having a recorded ICH.

### Respiratory viral exposures

Given changes in data collection, SARS-CoV-2 activity was represented by a composite series spanning two surveillance periods: test-adjusted case counts from Ontario’s Case and Contact Management System from March 2020 through August 2022, corrected for differential testing intensity using a previously described method (19–21), and percentage test positivity from the Respiratory Virus Detection Surveillance System (RVDSS) from September 2022 through March 2024 (22, 23). Combined influenza A/B and respiratory syncytial virus (RSV) activity were obtained from RVDSS for the full study period.

Pre-specified analyses focused on SARS-CoV-2, influenza A/B viruses, and RSV, but we also conducted exploratory analyses for five additional respiratory-virus groups (human parainfluenza virus, human metapneumovirus, adenovirus, enterovirus/rhinovirus, and seasonal coronaviruses) for which data were available from September 2015 onward, and for which time series were obtained after our primary pre- specified analysis had been completed. To express associations on a common scale while preserving a meaningful zero, each exposure was divided by its SD over the period in which its surveillance metric was used, without mean-centring; the two components of the SARS-CoV-2 series were scaled before concatenation. Zero therefore continued to represent no detected activity. We indexed and merged exposure, outcome, and population series by calendar week using one-to-one date keys and checked for duplicate dates and unmatched weeks.

### Statistical analysis

We first assessed whether mycobacterial presentations during the pandemic differed from the numbers expected from pre-pandemic patterns. Negative-binomial models with Fourier terms for seasonality and a population offset were fitted to 468 available pre-pandemic weeks. We used three representations of the underlying time trend (no trend, a linear trend, or a natural spline with 3 degrees of freedom) and projected expected weekly counts through the pandemic period. For each model, an intercept-only negative-binomial model with the log expected count as an offset estimated the observed-to-expected rate ratio. We also calculated cumulative excess presentations by summing weekly observed minus expected counts from March 15, 2020, onward.

As a complementary analysis, we used all 676 available weeks and estimated an average pandemic-era change while adjusting for the same seasonal and time-trend terms. Model fit was compared using the Akaike and Bayesian information criteria within each approach and outcome, but all six estimates, three from the projection approach and three from the full-series approach, were reported. Confidence intervals from the projection analysis treat expected counts as fixed and therefore omit some uncertainty about what would have happened in the absence of the pandemic. We therefore considered whether findings were consistent across plausible trend assumptions rather than relying on any one model.

We next modelled temporal associations between viral activity and mycobacterial presentations using distributed lag non-linear models (DLNM) with a linear exposure-response function and a natural-spline lag function with 3 degrees of freedom over a primary 0–12-week window (24). Models used quasi-Poisson errors, a population offset, first- and second-order Fourier terms for seasonality, and a natural spline for continuous weekly time with 13 degrees of freedom over the 13-year series. We report cumulative rate ratios per 1-SD increase in viral activity over the full lag window, together with lag-specific estimates. Sensitivity analyses extended the maximum lag for SARS-CoV-2 to 26 and 52 weeks.

We repeated the primary SARS-CoV-2 analysis by age (<65 and ≥65 years) and recorded ICH status and used Wald tests to assess between-stratum heterogeneity. Influenza and RSV were analysed using the same specification; all non-SARS-CoV-2 models were adjusted for concurrent SARS-CoV-2 activity. The five additional respiratory-virus groups were analysed over the shorter period for which they were available. These models assess whether weekly rises and falls in viral activity were followed by changes in mycobacterial presentations after accounting for seasonality and long-term trends. Unlike the pre-pandemic projection analyses, they do not require estimating expected counts in the absence of the pandemic.

The three prespecified viral exposures (SARS-CoV-2, influenza, and RSV) and two outcomes produced six primary virus-outcome comparisons. The five additional virus groups and two outcomes produced ten exploratory comparisons. We report two-sided nominal p values and 95% confidence intervals and indicate associations meeting Bonferroni thresholds within the primary exposure set (p<0.0083) and across all 16 comparisons (p<0.0031). Minimum detectable effects at 80% power were calculated from observed standard errors. Data preparation used Stata version 19; modelling used R with the dlnm and MASS packages within the CIHI Secure Access Environment.

### Ethical Approval

This study used de-identified administrative data in the CIHI Secure Access Environment and was approved by the Research Ethics Board of the University of Toronto (Protocol # 41690). Applications for access to CIHI data can be completed at https://www.cihi.ca/en/access-data-and-reports/make-a-data-request.

## RESULTS

### Descriptive epidemiology

Over 13 years, we identified 3,985 first TB-coded and 5,214 first NTM-coded acute-care presentations, corresponding to crude rates of 2.86 and 3.75 per 100,000 person-years, respectively (**Table 1**). Recorded ICH was present in 6.3% of TB and 13.5% of NTM presentations. Adults aged 65 years or older had higher adjusted rates of both outcomes than those younger than 20 years (TB rate ratio 2.81; NTM rate ratio 5.87), and male sex was associated with TB but not NTM. Crude pandemic-era rates were lower than pre-pandemic rates for both TB (2.70 vs 2.96 per 100,000 person-years) and NTM (3.26 vs 4.03).

**Table 1.** Descriptive epidemiology, tuberculosis and non-tuberculous mycobacterial disease, Ontario, 2011–2024.

| **Characteristic** | **Tuberculosis** | **Non-tuberculous mycobacteria** |
|---|---|---|
| Total presentations, n | 3,985 | 5,214 |
| Overall crude rate, per 100,000/year | 2.86 | 3.75 |
| Pre-pandemic crude rate | 2.96 | 4.03 |
| Pandemic crude rate | 2.70 | 3.26 |
| Immunocompromising host condition, % | 6.3% | 13.5% |
| Age 20–64 y (ref: <20 y) | 1.01 (0.88–1.16) | 0.81 (0.71–0.93) |
| Age ≥65 y (ref: <20 y) | 2.81 (2.44–3.24) | 5.87 (5.12–6.73) |
| Male sex (ref: female) | 1.31 (1.23–1.40) | 1.05 (0.99–1.11) |

**NOTE:** Rate ratios are adjusted for age, sex, Fourier seasonal terms, and secular trend. Outcomes are first TB- or NTM-coded acute-care presentations. ICH = recorded immunocompromising host condition.

### Pandemic-era changes in presentations

Estimates of pandemic-era change depended on how expected counts in the absence of the pandemic were modelled, so we report both pre-pandemic projection and full-series analyses (**Table 2**; **Figure 2**). Under the three projection models, TB rate ratios were 1.095 (1.037–1.156) with seasonality alone, 1.067 (1.010–1.127) with a linear trend, and 1.031 (0.976–1.089) with a 3-degree-of-freedom natural spline. Under the corresponding full-series models, estimates were 1.095 (1.022–1.174), 1.074 (0.955– 1.209), and 1.217 (1.035–1.431). All six point estimates exceeded unity, four confidence intervals excluded unity, and the estimated increase ranged from 3% to 22%.

**Figure 1.**
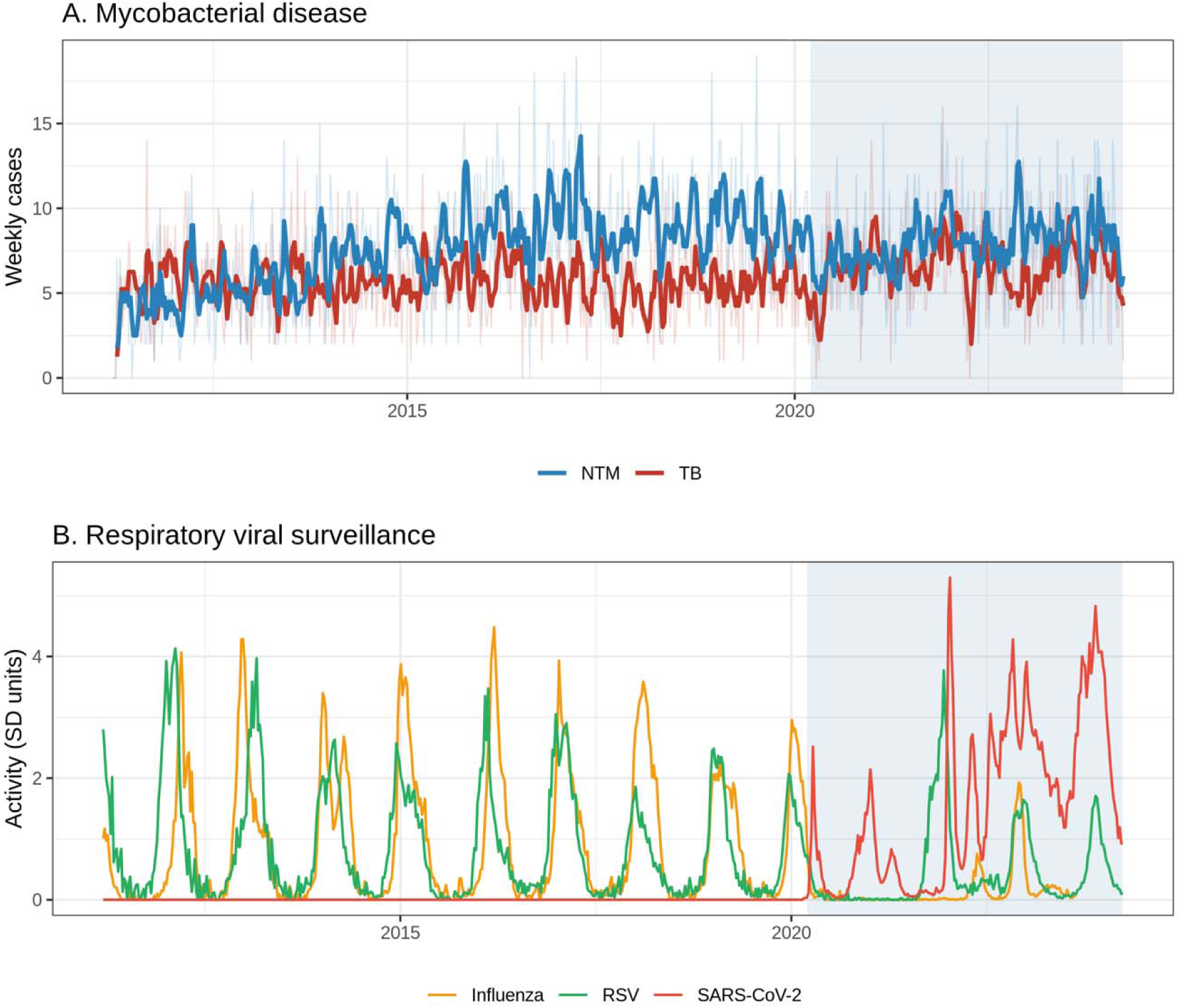
Temporal trends in mycobacterial acute-care presentations and respiratory-virus activity, Ontario, 2011-2024. (A) Weekly first TB- and NTM-coded acute-care presentations (bold lines, 4-week rolling means; faint lines, raw weekly counts). (B) Weekly SARS-CoV-2, influenza A, and RSV surveillance activity in SD units. The pandemic era is shaded from March 15, 2020.

**Figure 2.**
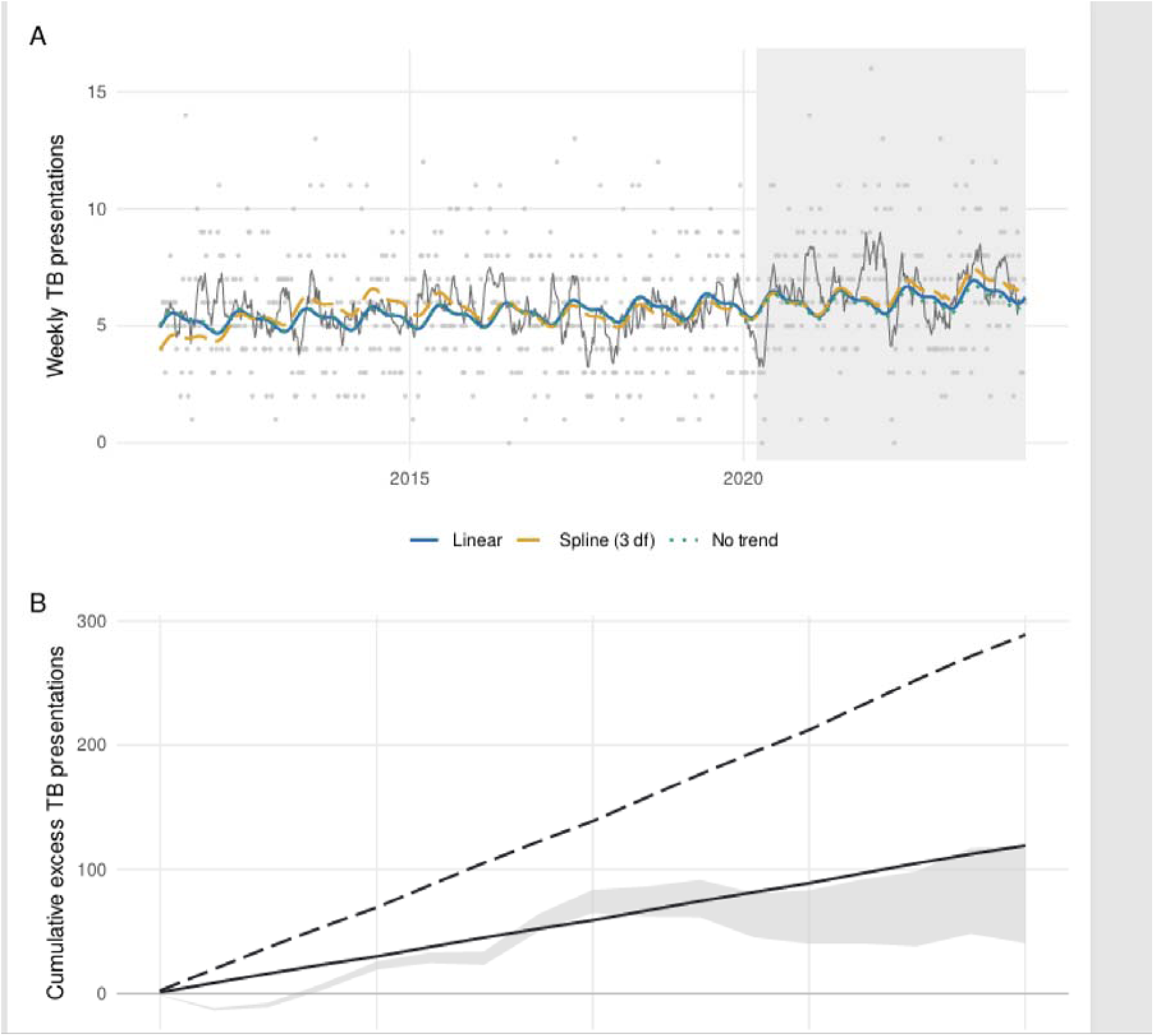
Observed TB-coded acute-care presentations, modelled expected counts, and cumulative excess, GTHA+, 2011–2024. (A) Grey points show weekly presentations and the dark-grey line an 8-week rolling mean. Coloured lines show expected counts under three assumptions about how the pre-pandemic trend would have continued after March 15, 2020. (B) Observed cumulative excess TB presentations, calculated as the running sum of observed minus expected counts from March 15, 2020 onward. The grey band spans the three projection models and therefore shows the range of observed excess consistent with alternative assumptions about the underlying trend. The two straight reference lines are not model fits: they show the excess that would have accumulated had presentations run at a constant 9.5% (solid line) or 21.7% (dashed line) above expectation throughout the pandemic era, these being the increases implied by the models favoured by the Bayesian and Akaike information criteria respectively. The observed excess accumulates through early 2022 and then flattens, whereas a constant proportional increase would continue to accumulate linearly, which is why an average pandemic-era rate ratio should not be read as a constant effect.

**Table 2.** Pandemic-era changes under alternative assumptions about underlying time trends, by outcome and estimation approach.

| **Approach and trend specification** | **TB rate ratio (95% CI)** | **AIC** | **BIC** | **NTM rate ratio (95% CI)** | **AIC** | **BIC** |
|---|---|---|---|---|---|---|
| ***Pre-pandemic fit, projected forward*** |  |  |  |  |  |  |
| Seasonality only, no trend | 1.095 (1.037–1.156) | 2254.0 | **2279.1** | 0.973 (0.924–1.025) | 2572.7 | 2597.7 |
| Linear in time | 1.067 (1.010–1.127) | 2255.8 | 2285.0 | 0.581 (0.552–0.612) | 2470.2 | 2499.4 |
| Natural spline, 3 df | 1.031 (0.976–1.089) | **2243.4** | 2280.9 | 0.959 (0.910–1.011) | **2401.5** | **2439.0** |
| ***Pandemic indicator, full series*** |  |  |  |  |  |  |
| Seasonality only, no trend | 1.095 (1.022–1.174) | 3253.7 | **3285.4** | 0.972 (0.905–1.045) | 3633.1 | 3664.8 |
| Linear in time | 1.074 (0.955–1.209) | 3255.6 | 3291.8 | 0.599 (0.537–0.667) | 3524.1 | 3560.3 |
| Natural spline, 3 df | 1.217 (1.035–1.431) | **3248.5** | 3293.9 | 0.849 (0.735–0.980) | **3454.2** | **3499.5** |

**NOTE:** Bold identifies the model favoured by each criterion within an approach and outcome. Projection models were fitted to the 468 pre-pandemic weeks and used to estimate expected pandemic-era counts; rate ratios compare observed with expected counts. Their confidence intervals treat expected counts as fixed and therefore do not include all uncertainty in the no-pandemic scenario. Full-series models used all 676 weeks and estimated an average pandemic-era change while adjusting for trend and seasonality. Information criteria can be compared only within the same approach and outcome.

Model-selection criteria did not identify a single preferred assumption. The Bayesian information criterion favoured the no-trend models, both of which estimated a rate ratio of 1.095, while the Akaike information criterion favoured the spline models, which estimated 1.031 under projection and 1.217 in the full-series analysis.

Depending on the model, the estimates implied approximately 40–280 excess TB-coded presentations over four years. Under the projection models, cumulative excess increased through early 2022 and then levelled off, ending between 41 and 119 cumulative excess presentations (**Figure 2B**). This pattern was not consistent with a constant increase throughout the four-year pandemic period.

For NTM, all six pandemic-era estimates were at or below unity. Projection-model rate ratios were 0.973 (0.924–1.025) with seasonality alone, 0.581 (0.552–0.612) with a linear trend, and 0.959 (0.910–1.011) with a 3-degree-of-freedom natural spline; the corresponding full-series estimates were 0.972 (0.905–1.045), 0.599 (0.537–0.667), and 0.849 (0.735–0.980). The linear models produced the largest apparent deficits because they continued the steep pre-pandemic increase into the pandemic era. No model suggested an increase, but the size of any decrease was uncertain.

### SARS-CoV-2 activity and mycobacterial presentations

Cumulative SARS-CoV-2 activity over 0–12 weeks was not positively associated with TB-coded presentations (cumulative rate ratio per 1-SD increase 0.909, 95% CI 0.822–1.006) or NTM-coded presentations (0.953, 0.868–1.047) (**Figure 3**; **Supplementary Table S1**). No lag-specific estimate showed a statistically significant positive association. Extending the maximum lag to 26 or 52 weeks did not reveal a positive cumulative association for either outcome (at 26 weeks, TB 0.897, 95% CI 0.746–1.078; NTM 0.862, 0.726–1.023; at 52 weeks, TB 0.639, 95% CI 0.318–1.281; NTM 1.233, 0.649–2.344).

**Figure 3.**
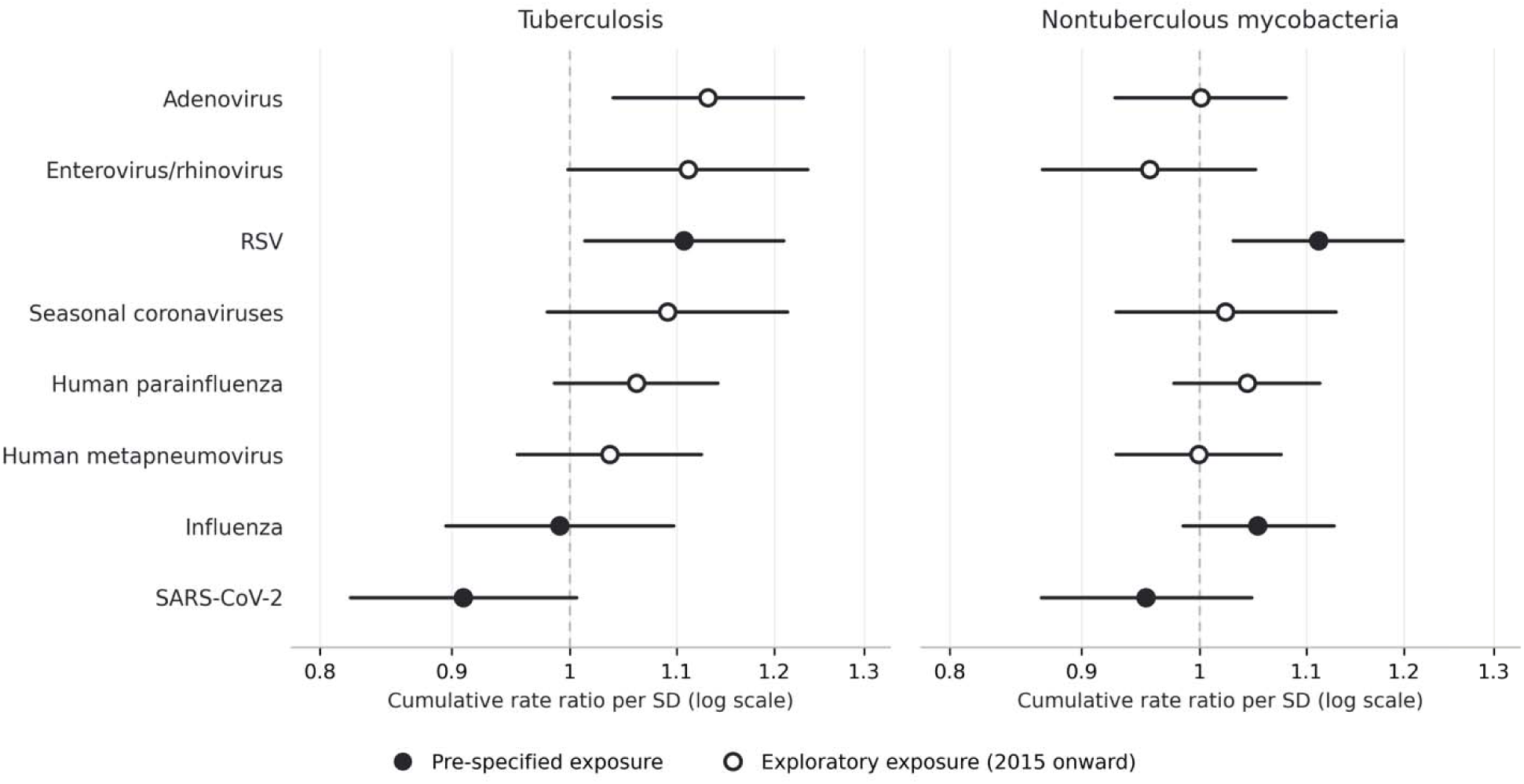
Cumulative rate ratios for TB- and NTM-coded acute-care presentations by respiratory-virus exposure. Forest plot of cumulative rate ratios per 1-SD increase in activity over a 0-12-week lag window, with 95% confidence intervals. All non-SARS-CoV-2 estimates, both prespecified and exploratory, are adjusted for concurrent SARS-CoV-2 activity. Filled circles denote the six primary comparisons; open circles denote exploratory comparisons involving the five additional virus groups available from 2015 onward. Exposures are ordered by the TB point estimate.

Age- and recorded-ICH–stratified estimates were similar to the overall results, with no evidence of heterogeneity by age (TB p=0.941; NTM p=0.787) or recorded ICH status (TB p=0.776; NTM p=0.072) (**Supplementary Table S1**). Although the NTM estimate in the recorded-ICH stratum excluded unity (cumulative rate ratio 0.754, 95% CI 0.578–0.984), this stratum included relatively few weekly presentations and the between-stratum test for heterogeneity was not significant.

### Comparator respiratory-virus exposures

Influenza activity was not associated with either outcome (TB cumulative rate ratio 0.991, 95% CI 0.895–1.097; NTM 1.053, 0.985–1.127), whereas RSV was positively associated with both TB (1.107, 1.013–1.210) and NTM (1.112, 1.030–1.199) presentations (**Figure 3**; **Supplementary Table S2**). The RSV–NTM association survived Bonferroni correction within the six primary virus–outcome comparisons (p=0.006), while RSV–TB was nominally significant (p=0.025). Among the five exploratory virus groups, only adenovirus was nominally associated with TB (rate ratio 1.131, 95% CI 1.039–1.231; p=0.005); no exploratory association met the experiment-wide Bonferroni threshold (**Supplementary Table S3**).

## DISCUSSION

In this population-based study, TB-coded acute-care presentations were probably modestly elevated during the pandemic era, but we could not attribute these increases to direct effects of SARS-CoV-2 activity, at lags up to 52 weeks. The estimated magnitude of the pandemic-era TB increase depended on how we represented what would have happened without the pandemic. Models that projected pre-pandemic patterns beyond 2020 and models that estimated an average pandemic-era change from the full series produced estimates of different magnitudes because they made different assumptions about the underlying trend in cases. A flexible trend can also behave differently when extended beyond 2020 than when fitted to the entire series, which is why our two approachs to estimating the effect of the pandemic on disease trends diverge most where splines were included in our models. However, the consistent direction across all models supports an increase, whereas the 3%-22% range in this increase conveys the uncertainty more clearly than any single selected estimate.

We evaluated SARS-CoV-2 effects on mycobacterial disease risk at a population level by treating viral activity as an ecological exposure. This approach has the advantage of not being subject to biases related to differential SARS-CoV-2 testing probability in individuals with medical comorbidity. However, population- and individual-level relative risks may be quite different in magnitude, even for an identical individual-level effect. A population time-series estimate applies to the entire population, although only a fraction of that population has been recently infected. A large individual-level effect confined to a small subgroup may therefore produce only a modest population-level association. We recently demonstrated this distinction for pulmonary embolism: an Ontario time-series analysis found a 12% increase per SD of SARS-CoV-2 activity, whereas a Swedish individual-level study reported a greater than 30-fold increase following infection (25, 26).

A similar dilution could occur for TB if elevated risk were concentrated among people with severe COVID-19 or particular vulnerabilities. A nationwide Chilean cohort of 3.6 million adults, excluding those with recognized immunocompromise, found that severe COVID-19 was associated with a markedly increased risk of incident TB that peaked within the first six months and was attenuated by prior SARS-CoV-2 vaccination (27). Baseline latent and subclinical TB status was unavailable, however, and the study could not distinguish effects of severe infection from those of treatment, detection, or underlying vulnerability.

In our primary 12-week analysis, we had 80% power to detect cumulative rate ratios of approximately 1.16 for TB and 1.14 for NTM per SD of activity. The null estimates we identified therefore argue against a large population-level effect but do not exclude a clinically important individual-level effect in a smaller subgroup. The TB point estimate below 1 should not be interpreted as protective, but an incidence rate ratio below 1 may suggest that viral surges could have delayed TB diagnosis and attenuated a short-term association. Analyses extending the maximum lag to 26 and 52 weeks also detected no association, arguing against a delayed effect at the population level.

Influenza and RSV were included in our analysis *a priori* to assess whether an anticipated SARS-CoV-2 association was virus-specific or reflected patterns shared across respiratory-virus seasons. Contrary to this expectation, SARS-CoV-2 and influenza were not positively associated with either outcome, whereas RSV was positively associated with TB and NTM; adenovirus was also nominally associated with TB. Only the RSV-NTM association remained after Bonferroni correction. Correlated seasonality, changes in healthcare contact, and diagnostic opportunity could produce non-causal associations. However, respiratory viruses can also have important immunological effects, and genuine virus-specific effects on mycobacterial disease are biologically plausible (28), and our ecological design cannot distinguish these possibilities. The comparison is also not symmetric in precision: influenza and RSV were observed over the full thirteen years and across many epidemic cycles, whereas the SARS-CoV-2 series spans only the four years from 2020 and a smaller number of distinct waves, so the prespecified comparators were estimated from considerably more information. Rather than providing the anticipated specificity test for SARS-CoV-2, these findings generate hypotheses concerning other respiratory viruses that require investigation using individual-level data.

While we identify a pandemic-era TB increase in central Ontario similar to that reported in other jurisdictions (1, 2, 5, 6), we are unable to resolve its cause. Changes in population composition and migration, disruptions to latent-TB testing and preventive treatment, delayed diagnosis, and altered healthcare use or ascertainment may all have contributed. NTM presentations did not show a corresponding increase, possibly because reductions in elective imaging, bronchoscopy, and respiratory cultures reduced ascertainment of pulmonary NTM early in the pandemic. A related possibility is that the ambulatory component of our outcome was differentially affected: pulmonary NTM is diagnosed in ambulatory settings more often than TB, and in-person ambulatory care contracted sharply in 2020 while virtual visits, which are not captured in the National Ambulatory Care Reporting System, expanded. Our data do not permit presentations to be separated by care setting, so we cannot quantify this contribution. To explore whether two candidate mechanisms could plausibly account for the TB excess, we developed an interactive scenario tool in which readers can vary literature-informed assumptions concerning disruption of latent-TB care and migration (https://fismanda.github.io/mycobacteria-and-respiratory-viruses/; archived at DOI: 10.5281/zenodo.22216420). The tool assesses order-of-magnitude plausibility rather than causal attribution.

Strengths of this study include its large, geographically concentrated population; long pre-pandemic baseline; separate evaluation of the pandemic-era increase and its hypothesized relationship with SARS-CoV-2 activity; sensitivity analyses extending the lag window to one year; and presentation of results under several assumptions about how pre-pandemic trends would have continued. Important limitations are the ecological nature of viral exposure, absence of individual data on infection severity, vaccination, migration history, and subclinical TB, and use of a composite SARS-CoV-2 exposure spanning two surveillance systems. Outcomes were first health care presentations identified through administrative codes rather than validated TB or NTM diagnoses or surveillance notifications, and because the ambulatory records available to us could not be separated into emergency and clinic encounters we could not restrict the outcome to unscheduled care, medication-induced immunosuppression could not be captured, and the size of immune compromised strata was small. NTM was analysed as a composite outcome despite comprising clinically heterogeneous conditions with different anatomical and immunological determinants.

In conclusion, TB-coded acute-care presentations appear to have increased modestly during the pandemic era in central Ontario, Canada, but community SARS-CoV-2 activity was not positively associated with subsequent TB or NTM presentations over lag windows from 12 weeks to one year, and did not appear to explain the population-level TB increase. Clinically important individual-level effects remain possible, particularly after severe COVID-19, and would require studies linking individual infection histories, migration, and clinical data with validated TB outcomes.

## SUPPLEMENTARY TABLES

**Supplementary Table S1.**
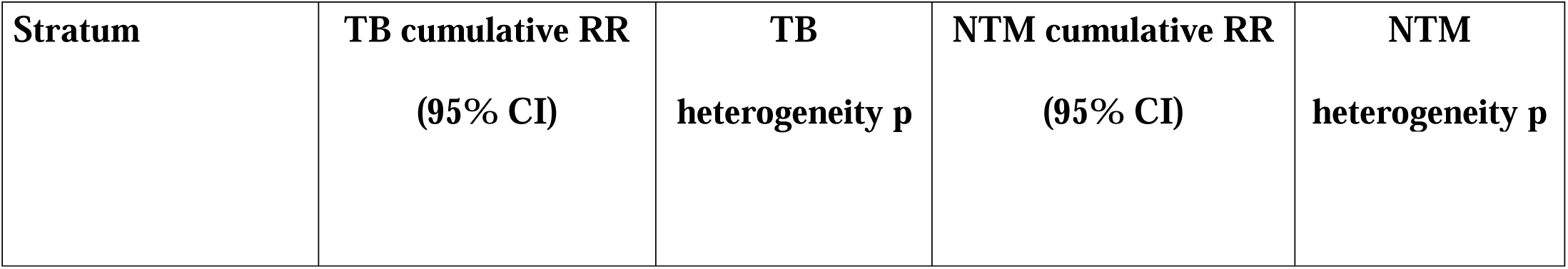
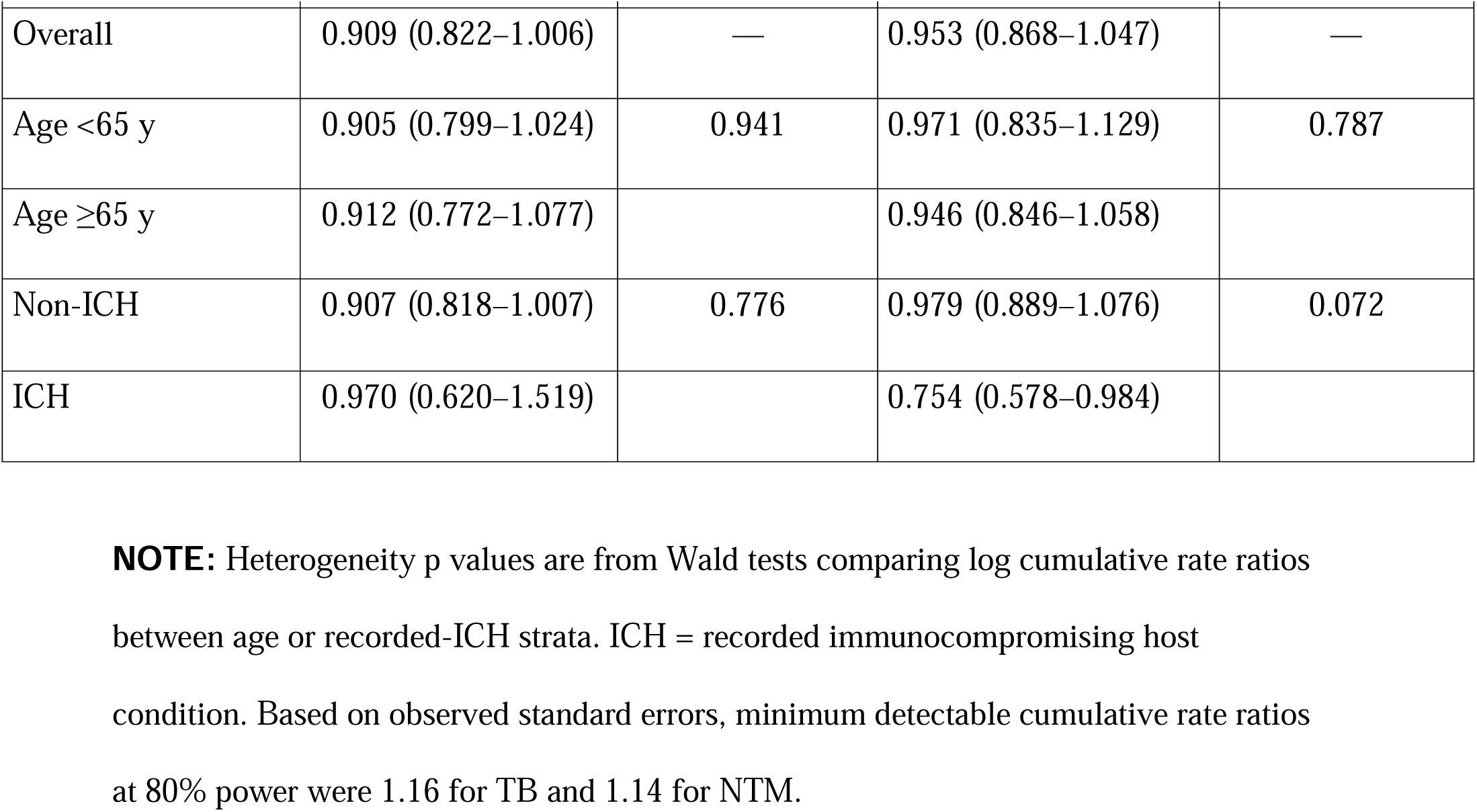
Cumulative rate ratios for TB- and NTM-coded acute-care presentations associated with SARS-CoV-2 activity (per 1-SD increase; 0–12-week lag), overall and by age and recorded immunocompromising condition.

**Supplementary Table S2.**
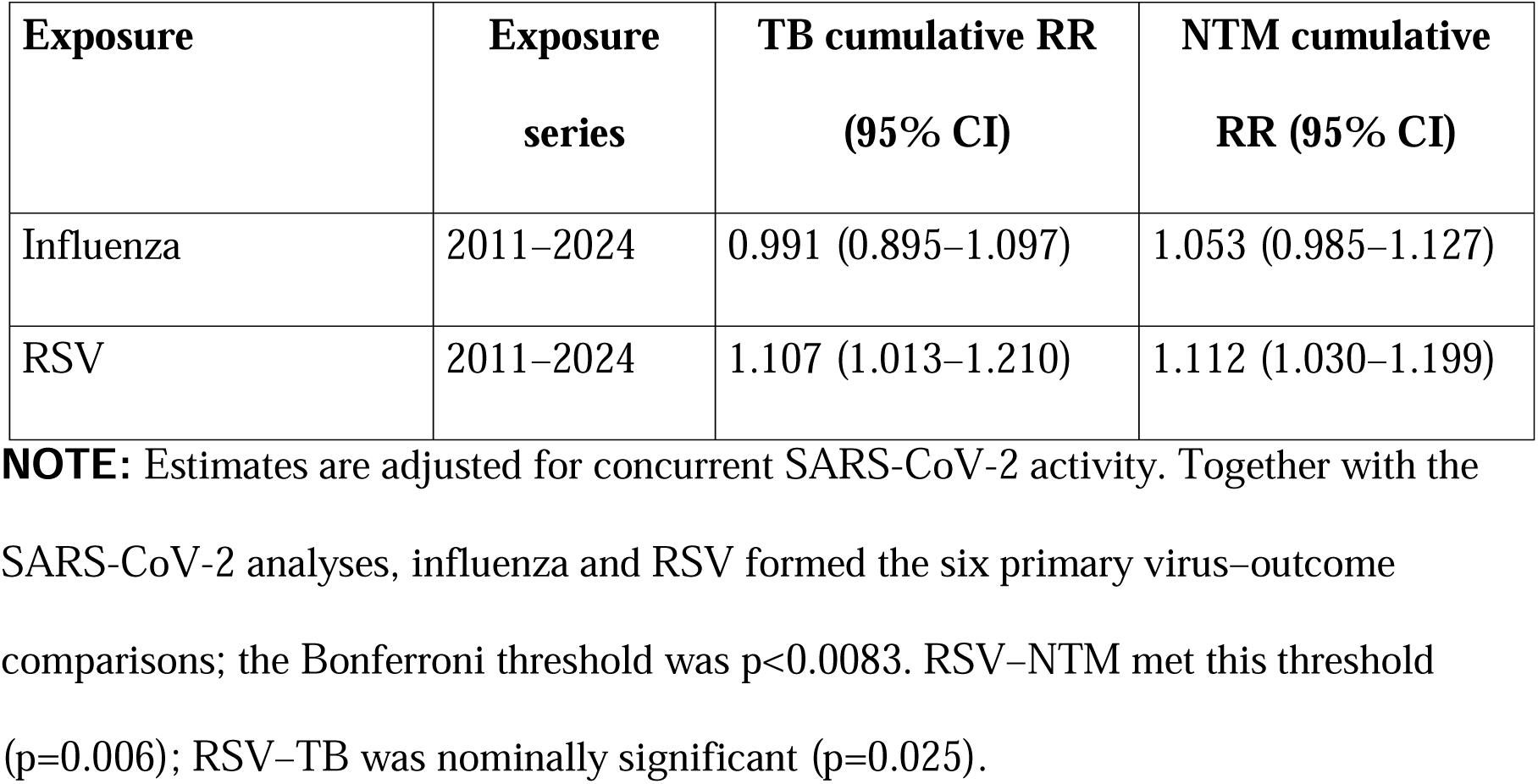
Cumulative rate ratios for TB- and NTM-coded acute-care presentations associated with prespecified non-SARS-CoV-2 respiratory-virus exposures (per 1-SD increase; 0–12-week lag).

| **Exposure** | **Exposure series** | **TB cumulative RR (95% CI)** | **NTM cumulative RR (95% CI)** |
|---|---|---|---|
| Influenza | 2011–2024 | 0.991 (0.895–1.097) | 1.053 (0.985–1.127) |
| RSV | 2011–2024 | 1.107 (1.013–1.210) | 1.112 (1.030–1.199) |

**NOTE:** Estimates are adjusted for concurrent SARS-CoV-2 activity. Together with the SARS-CoV-2 analyses, influenza and RSV formed the six primary virus–outcome comparisons; the Bonferroni threshold was p<0.0083. RSV–NTM met this threshold (p=0.006); RSV–TB was nominally significant (p=0.025).

**Supplementary Table S3.** Cumulative rate ratios for TB- and NTM-coded acute-care presentations associated with exploratory respiratory-virus exposures (per 1-SD increase; 0–12-week lag).

| **Exposure** | **TB cumulative RR (95% CI)** | **NTM cumulative RR (95% CI)** |
|---|---|---|
| Human parainfluenza virus | 1.061 (0.986–1.141) | 1.043 (0.977–1.113) |
| Human metapneumovirus | 1.036 (0.954–1.124) | 0.999 (0.928–1.075) |
| Adenovirus | 1.131 (1.039–1.231) | 1.001 (0.927–1.080) |
| Enterovirus/rhinovirus | 1.111 (0.998–1.236) | 0.956 (0.869–1.051) |
| Seasonal coronaviruses | 1.091 (0.980–1.214) | 1.023 (0.928–1.129) |

**NOTE:** Estimates are adjusted for concurrent SARS-CoV-2 activity and use the shorter period for which all five exploratory virus series were available (September 2015–March 2024). These analyses were exploratory. Adenovirus–TB was nominally significant (p=0.005); no exploratory association met the experiment-wide Bonferroni threshold of p<0.0031 across all 16 virus–outcome comparisons.

## SUPPLEMENTARY APPENDIX: STUDY REGION DEFINITION AND POPULATION CHARACTERISTICS

### Geographic coverage

The Greater Toronto–Hamilton Area Plus (GTHA+) comprises 14 Public Health Units (PHUs) in central Ontario, Canada (**Figure S1**). The region consists of the 6 PHUs of the Greater Toronto–Hamilton Area (Toronto, Hamilton, Peel, York, Durham, and Halton), plus 8 bordering PHUs: Simcoe-Muskoka, Haliburton–Kawartha–Pine Ridge, Peterborough, Wellington–Dufferin–Guelph, Waterloo, Brant, Niagara, and Haldimand-Norfolk.

### Rationale for study region

The GTHA+ was selected to provide a large population concentrated in a relatively small geographic area, improving the relevance of common ecological exposure series. The region represents approximately 70% of Ontario’s population and 27% of Canada’s population while comprising about 5% of Ontario’s land area. Its concentration of healthcare institutions also improves statistical precision for acute-care outcomes.

### Population characteristics

Population estimates for each PHU were obtained from Statistics Canada, which provides annual figures for both census and non-census years. As of 2024, the GTHA+ had an estimated population of 11.2 million residents. The region experienced substantial growth during the study period, with overall population increasing 41.3% between 2001 and 2024 (**Figure S2**). Growth rates varied across constituent PHUs, ranging from 18.6% (Haldimand-Norfolk) to 61.1% (Peel).

**Figure S1.**
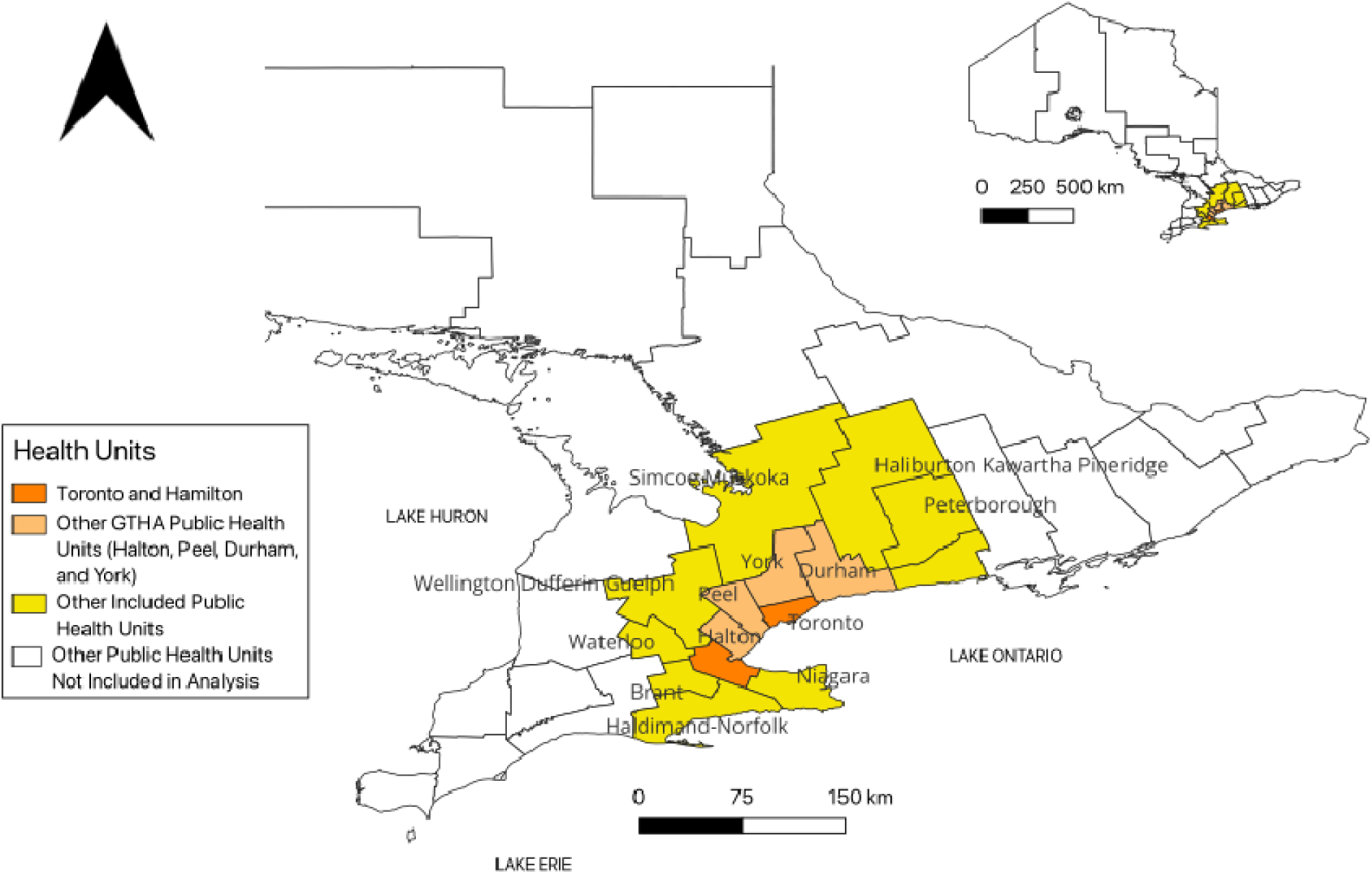
Map of Public Health Units included in the GTHA+ study region. The main map shows the 14 Public Health Units comprising the GTHA+ region within southern Ontario. Toronto and Hamilton are shown in dark orange; other GTHA Public Health Units (Halton, Peel, Durham, York) in light orange; and the 8 additional PHUs in yellow. Public Health Units not included are shown in white. The inset map shows the location of the GTHA+ region within Ontario. Scale bars indicate 0–150 km (main) and 0–500 km (inset).

**Figure S2.**
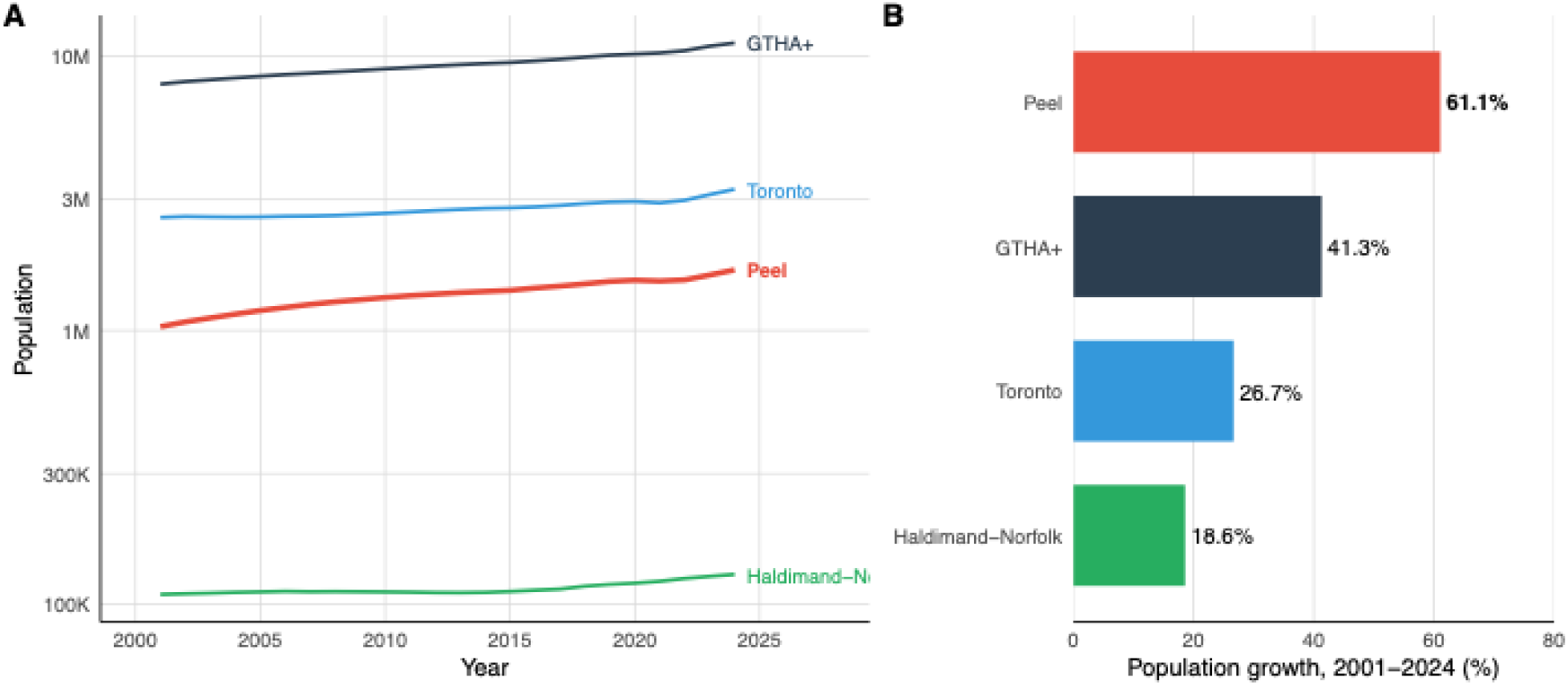
Population growth in the GTHA+ region, 2001–2024. Panel A shows population trajectories on a log scale for the overall GTHA+ region and for the largest (Toronto), smallest (Haldimand-Norfolk), and most rapidly growing (Peel) Public Health Units. Panel B shows percent population growth over the study period for these units.

## ACKNOWLEDGEMENTS

The authors thank the Canadian Institute for Health Information (CIHI) for providing access to the Discharge Abstract Database and National Ambulatory Care Reporting System through the Secure Access Environment, and the Public Health Agency of Canada for the Respiratory Virus Detection Surveillance System data. The analyses, conclusions, opinions, and statements expressed herein are solely those of the authors and do not reflect those of the data sources; no endorsement is intended or should be inferred.

## Declaration of generative AI and AI-assisted technologies in the manuscript preparation process

During the preparation of this work the authors used Claude (Anthropic) and ChatGPT (OpenAI) for statistical code review and editorial revision. After using these tools, the authors reviewed and edited the content as needed and take full responsibility for the content of the published article.

## Data Availability

Analysis code is available at DOI: 10.5281/zenodo.22216420. The companion interactive scenario tool runs in any browser at https://fismanda.github.io/mycobacteria-and-respiratory-viruses/ and is archived at DOI: 10.5281/zenodo.22216420. Individual-level administrative health data were accessed within the Canadian Institute for Health Information Secure Access Environment. Respiratory-virus surveillance data are publicly available through the Public Health Agency of Canada.

